# Pandemic ST131 *Escherichia coli* presenting the UPEC/EAEC and ExPEC/EAEC hybrid pathotypes recovered from extraintestinal infections in a clinical setting of the Brazilian Amazon region

**DOI:** 10.1101/2024.03.11.24304096

**Authors:** Nathália M. S. Bighi, Érica L. Fonseca, Fernanda S. Freitas, Sérgio M. Morgado, Ana Carolina P. Vicente

**Affiliations:** Laboratório de Genética Molecular de Microrganismos, Instituto Oswaldo Cruz, FIOCRUZ, Rio de Janeiro, Brazil

**Keywords:** HyPEC, ExPEC, ST69, ST131, virulence marker, pandemic lineage

## Abstract

*Escherichia coli* is part of the commensal microbiota of human’s and animal’s gut. However, they may become pathogenic due to the acquisition of virulence factors that provide the ability to cause intestinal or extraintestinal infections, which makes *E. coli* the main cause of diarrheagenic diseases and urinary tract infections (UTIs) worldwide, respectively. Some strains, known as hybrids, may harbour a mix of virulence determinants of both diarrheagenic (DEC) and extraintestinal *E. coli* (ExPEC) pathotypes. Reports of hybrid *E. coli* in Brazil are rare, and the lineages associated with such pathotypes were poorly explored. This study aimed to characterize *E. coli* strains recovered from extraintestinal infections in a clinical setting of the Brazilian Amazon Region by means of lineage determination, antibiotic resistance profile, and investigation of DEC and ExPEC virulence markers. Fifteen ExPEC strains were recovered from distinct extraintestinal sites from inpatients of the General Hospital of Roraima (GHR), placed in the Brazilian Amazon region. Antibiotic susceptibility test revealed that all strains were multidrug-resistant and most of them, including those recovered from urine, were resistant to fluoroquinolones, the main therapeutic option for treating UTIs, probably due to the presence of Ser83Leu and Asp87Asn substitutions in GyrA. The MLST analysis revealed the polyclonal nature of these ExPEC strains since 11 STs were determined, including local and pandemic lineages, such as ST69 and ST131. Among the 15 isolates, 12 were classified as hybrids, due to the presence of the *aggR* virulence marker of the Enteroaggregative *E. coli* (EAEC) pathotype together with at least one ExPEC (*iutA, KPSMTII, sfaDE, papC, afaBC, iucD*) or Uropathogenic *E. coli* (UPEC) (*vat*, *fuyA*, *chuA* and *yfcV)* virulence determinants. These UPEC/EAEC (n=10) and ExPEC/EAEC (n=2) hybrid strains were found among distinct lineages, including new STs, and phylogroups (ST131/B2; ST1196/AxB1; ST9403/A; ST12394/A; NEW1-CC14/B2; NEW2-CC155/B1; NEW3-CC155/B1; NEW4-CC131/B2) and, for the first time, a hybrid phenotype was found in the pandemic ST131 lineage in Brazil. Therefore, this study provides new information on the epidemiological scenario of hybrid *E. coli* strains, contributing to a better understanding of the occurrence and pathogenic potential of these organisms.

## INTRODUCTION

*Escherichia coli* inhabits the intestinal tract of humans and other animals as an important member of their microbiota [1]. However, the acquisition by horizontal gene transfer of virulence determinants by certain *E. coli* clones has enabled them to cause not only intestinal but also extraintestinal infections in different hosts. In this way, based on the body site of infection, the host, and the presence of specific virulence markers, these bacteria can be classified into diarrheagenic (DEC), mainly featured by the presence of specific virulence factors directly related to diarrhoea, and extraintestinal pathogenic (ExPEC) *E. coli*, defined primarily by their site of isolation [2]. The DEC group encompasses several pathotypes, including enterotoxigenic (ETEC), enteropathogenic (EPEC), enteroinvasive (EIEC), enteroaggregative (EAEC), and Shiga toxin-producing (STEC) *E. coli*. EPEC and EAEC are considered the major pathotypes of diarrheagenic *E. coli* causing disease worldwide, including in Brazil. These pathotypes carry genetic determinants involved with disease development and can be used as molecular markers of the pathotype.

The ExPEC group includes the pathotypes neonatal meningitis-associated (NMEC), human sepsis-associated (SEPEC), avian pathogenic (APEC), and uropathogenic (UPEC) *E. coli*, which is the most prevalent pathotype among extraintestinal infections worldwide. UPEC is particularly associated with urinary tract infections (UTIs) and is a leading cause of both community-acquired and healthcare-associated UTIs. Currently, the most frequently reported high-risk ExPEC lineages belonging to the MLST sequence type ST131, ST69, ST10, ST405, ST38, ST95, ST648, ST73 and ST1193, which are known for their high prevalence in extraintestinal infections worldwide and for their significant role in global spread of multidrug resistance [3].

Studies *in vivo* revealed that the presence of at least two of five virulence genes (*pap*, *afa/dra*, *sfa*, *kpsMTII*, and *iut/iuc*) could be enough to cause extraintestinal infection in immunocompetent individuals [4], and that *E. coli* co-harbouring of *chuA*, *fyuA*, *vat*, and *yfcV* virulence genes were able to cause UTIs [5]. Additionally, due to horizontal gene transfer events, organisms may emerge as hybrid strains capable of causing intestinal and extraintestinal diseases due to the presence of virulence factors of both ExPEC and DEC pathotypes [6].

Strains harbouring a blend of virulence markers of different pathotypes have been described worldwide in both high-risk and local lineages, however, most of them correspond to hetero-pathogenic strains (characterized by a combination of virulence markers from DEC pathotypes) [7–9]. A recent study in South Africa demonstrated the high prevalence of several different DEC hybrid pathotypes, including strains sharing three virulence markers of three different DEC pathotypes, which were recovered from inanimate surfaces and presented a higher resistance profile compared to the classic pathotypes identified [8]. Another report demonstrated the occurrence of different DEC hybrid strains recovered from healthy individuals in Mexico [9].

In Brazil, few studies have characterized and reported the occurrence of hybrid strains in extraintestinal infection cases harbouring the combination of DEC and ExPEC or UPEC virulence markers, such as UPEC/EAEC, UPEC/aEPEC, ExPEC/STEC, ExPEC/aEPEC [10–18]. However, there is a lack of association concerning the STs of such hybrid strains circulating in the country. Moreover, most of these reports on hybrid strains were restricted to São Paulo, a cosmopolitan city of the Southeast Brazilian region, and until now, all studies reporting *E. coli* infections in the Amazon Region were associated only with diarrheagenic infections [19–22]. Therefore, further studies are required to investigate the distribution and potential impact of these strains on the clinical outcomes of affected individuals. For these reasons, the objective of this study was to characterize *E. coli* strains recovered from extraintestinal infections in the Brazilian Amazon Region by means of lineage determination (MLST), antibiotic resistance profile, and investigation of DEC and ExPEC virulence markers.

## METHODS

### Clinical data, bacterial strains and antimicrobial susceptibility test

From December 2016 to February, 2018, 15 *E. coli* isolates were recovered from extraintestinal infection cases at the General Hospital of Roraima (GHR), placed in Boa Vista. The strains were isolated from blood (n=2), urine (n=7), catheter tip (n=1), vagina secretion (n=1), leg secretion (n=1), soft tissue (n=1), and operative wound secretion (n=2). Species identification was performed with the automated VITEK2, and confirmed by sequencing the 16S rRNA and the MLST genes. These strains are part of the Bacterial Culture Collection of the Laboratory of Molecular Genetics of Microrganisms, FIOCRUZ.

### Antimicrobial Susceptibility testing

The antibiotic susceptibility profile was determined by disc-diffusion method according to clinical and laboratory standards institute (CLSI) guidelines [23] For the following antibiotics: gentamicin, amikacin, kanamycin, tobramycin, neomycin, streptomycin, imipenem, meropenem, ertapenem, cephalothin, cefoxitin, cefuroxime, ceftazidime, ceftriaxone, cefotaxime, cefepime, azithromycin, clarithromycin, erythromycin, ampicillin, ampicillin/sulbactam, amoxicillin, amoxicillin/clavulanic acid, carbenicillin, penicillin, aztreonam, piperacillin/tazobactam, ticarcillin/clavulanic acid, nalidixic acid, ciprofloxacin, norfloxacin, levofloxacin, ofloxacin, colistin sulphate, sulphonamide, trimethoprim, sulfamethoxazole/trimethoprim, chloramphenicol, fosfomycin, nitrofurantoin, tetracycline and minocycline.

### Genotyping by Multilocus sequence typing (MLST)

The genetic relationship among the ExPEC strains was established by MLST based on the Achtman MLST scheme (*adk, fumC, gyrB, icd, mdh, purA* and *recA*) (http://mlst.warwick.ac.uk). Clonal complexes (CCs) were considered when sequence types (STs) shared five or more identical alleles taking into account the seven genes considered in the MLST scheme.

### Detection of resistance genes by polymerase chain reaction (PCR) and sequencing

The isolates were screened by PCR and sequencing for the presence of the genes frequently associated with β-lactam resistance in *E. coli* such as *bla*_SHV_, *bla*_GES_, *bla*_TEM_, *bla*_CTX-M_ (class A β-lactamases). The plasmid-mediated fluoroquinolone (FQ) resistance *qnr* genes and the presence of mutations in the quinolone resistance-determining region (QRDR) of *gyrA* was also investigated (Table 1).

**Table 1.**
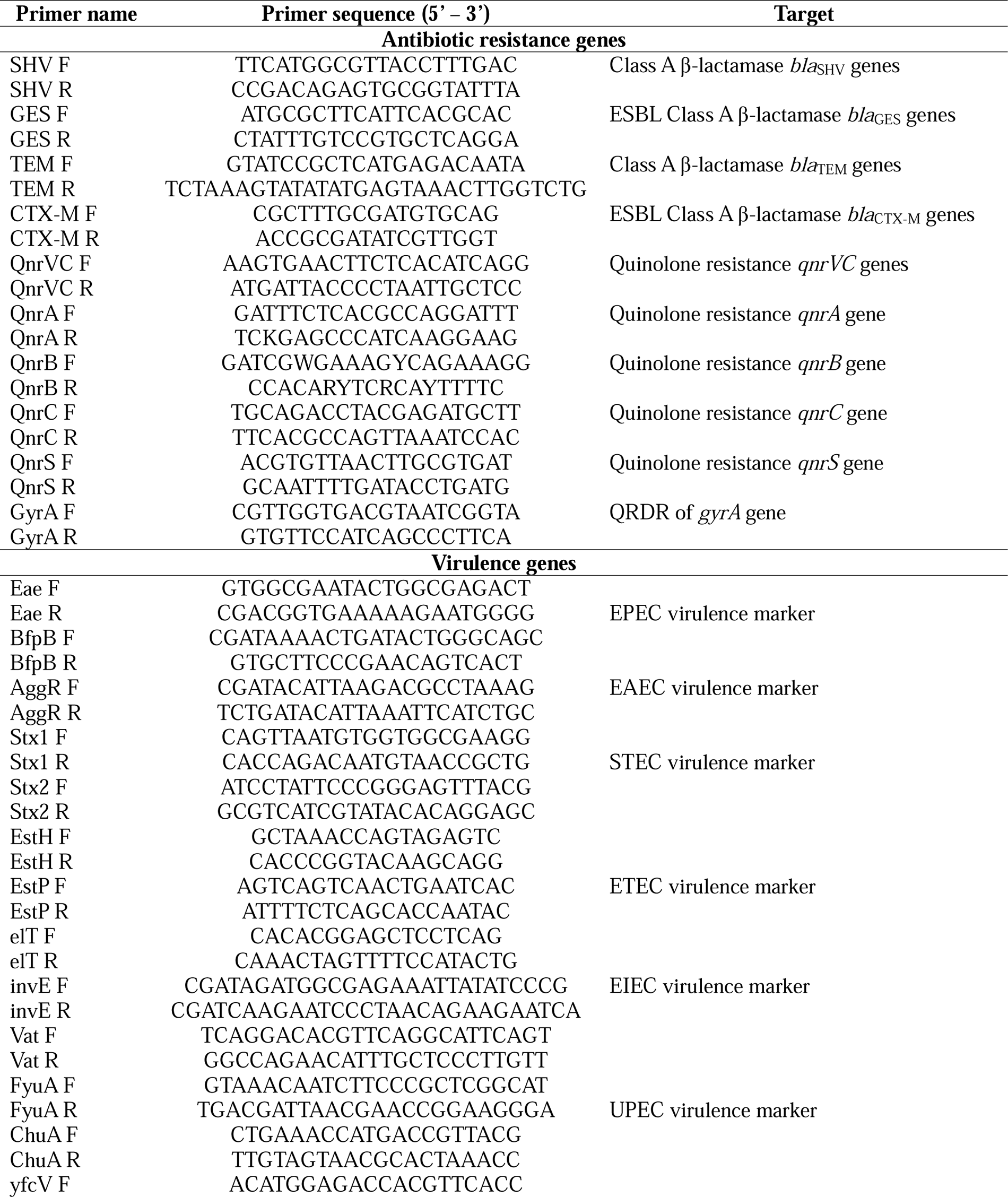
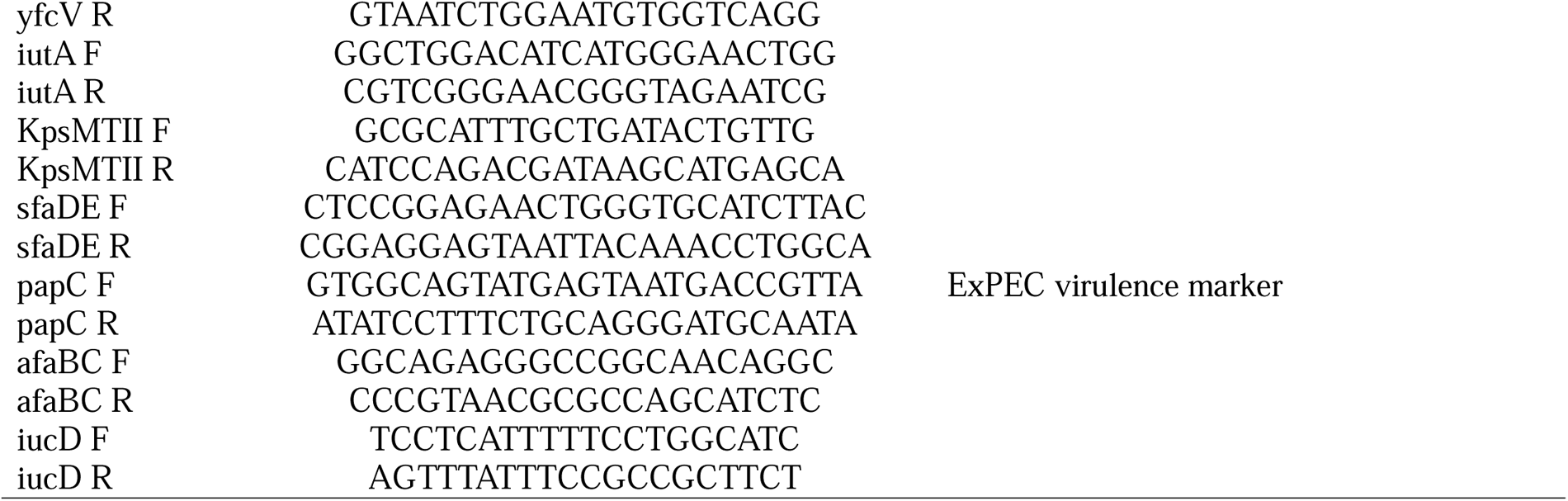
Primers used in this study.

### Molecular Characterization of Hybrid Strains

The presence of DEC and ExPEC virulence determinants was screened by PCR (Table 1). Those strains presenting at least one of the DEC pathotypes virulence markers were considered hybrid strains [13]. The DEC virulence factors screened were *eae* and *bfpB* genes, associated with epithelial adhesion in EPEC; the *aggR* gene, a universal regulator of EAEC virulence involved with the aggregative adhesion fimbriae (AAF) production; the *stx1* and *stx2* genes, related to the Shiga toxins production in STEC; the *estH*, *estP* and *elT*, involved with the production of heat-stable and heat-labile toxins in ETEC; the *invE* gene that encodes a regulatory protein that controls the transcriptional of invasion genes in EIEC pathotype; the *vat*, *fuyA*, *chuA* and *yfcV*, all markers of UPEC; and ExPEC markers *iutA*, *KPSMTII*, *sfaDE*, *papC*, *afaBC*, *iucD*.

## RESULTS AND DISCUSSION

The phenotypic analysis revealed that all *E. coli* ExPEC strains (n=15) were classified as multidrug-resistant (MDR) according to resistance classification criteria [24] (Table 1). Among the β-lactam antibiotics, all isolates were resistant to at least one of these antibiotics, and such phenotype could be explained by the presence of the *bla*_TEM_ gene in 14/15 isolates. In fact, *bla*_TEM_ is frequently associated with plasmids in *E. coli*, and it was the most prevalent gene among ExPEC recovered from UTI cases in São Paulo, Brazil [25]. However, although the *bla*_CTX-M_ has been frequently associated with ExPEC in Brazil and worldwide [26], this gene was not found in our sample, as well as the other screened β-lactamase genes *bla*_GES_ and *bla*_SHV_.

Moreover, a high prevalence of fluoroquinolone resistance was observed (73%) among the ExPEC strains. Although none of them carried any *qnr* allele, all FQ-resistant isolates presented mutations in the Quinolone Resistance-Determining Region (QRDR) of *gyrA* that led to the amino acid substitution Ser83Leu and Asp87Asn, known to be involved with FQ resistance emergence [27]. Interestingly, these substitutions were recently reported as the determinants of FQ resistance in ExPEC strains recovered from UTI cases in São Paulo [12].

The MLST analysis revealed a great diversity, in which 11 STs were assigned to the 15 ExPEC strains: ST69, ST131, ST8886, ST1196, ST9403, ST3180, ST12394, and four different new sequence types belonging to CC155 (n=2), CC14 (n=1) and CC131 (n=1), demonstrating the polyclonal nature of these hybrid strains. The most prevalent lineage among our sample was ST131, a pandemic clonal complex of the B2 phylogroup often associated with an arsenal of resistance and virulence genes, contributing to the success of this clone in spreading through clinical settings worldwide. The ST69 is another pandemic lineage belonging to the phylogroup D usually associated with clinics and food/environment contamination in several countries [27]. As found here, Lara and Colleagues have demonstrated the occurrence of ExPEC ST69 presenting the hybrid pathotype UPEC/EAEC in Brazil [10]. However, in spite of ST131 prevalence in Brazil, this is the first report of this lineage presenting a hybrid pathotype (UPEC/EAEC) in that country (Table 2).

**Table 2.**
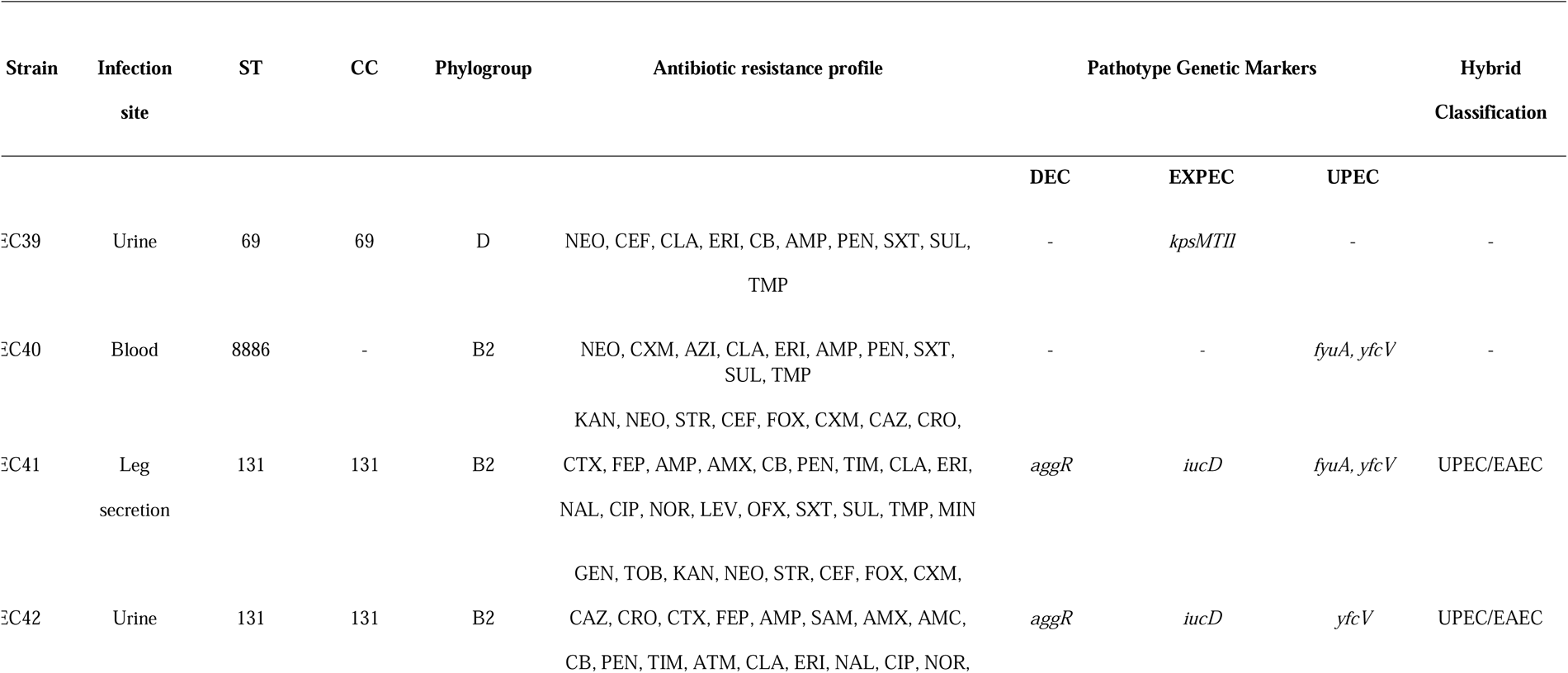
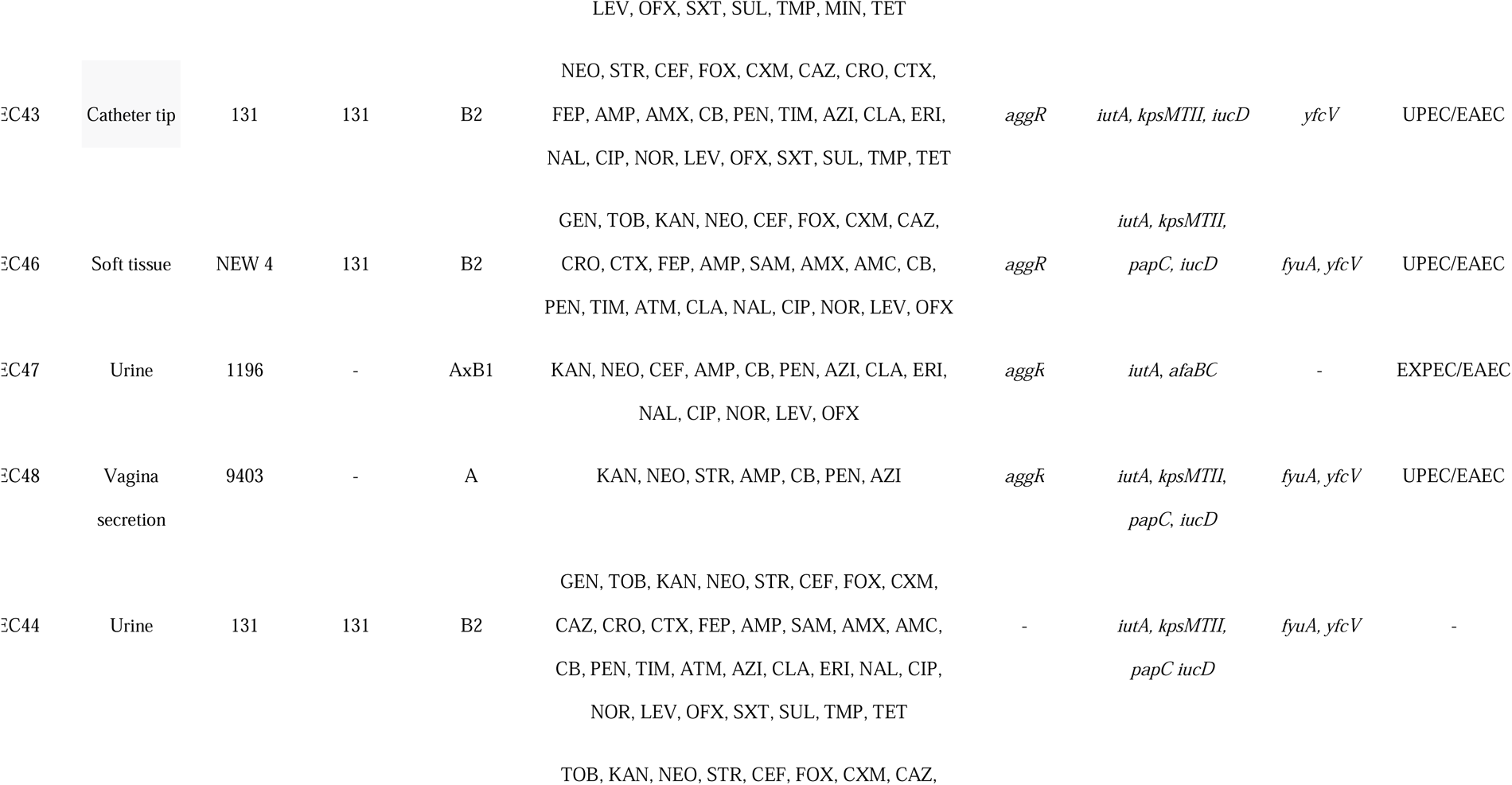
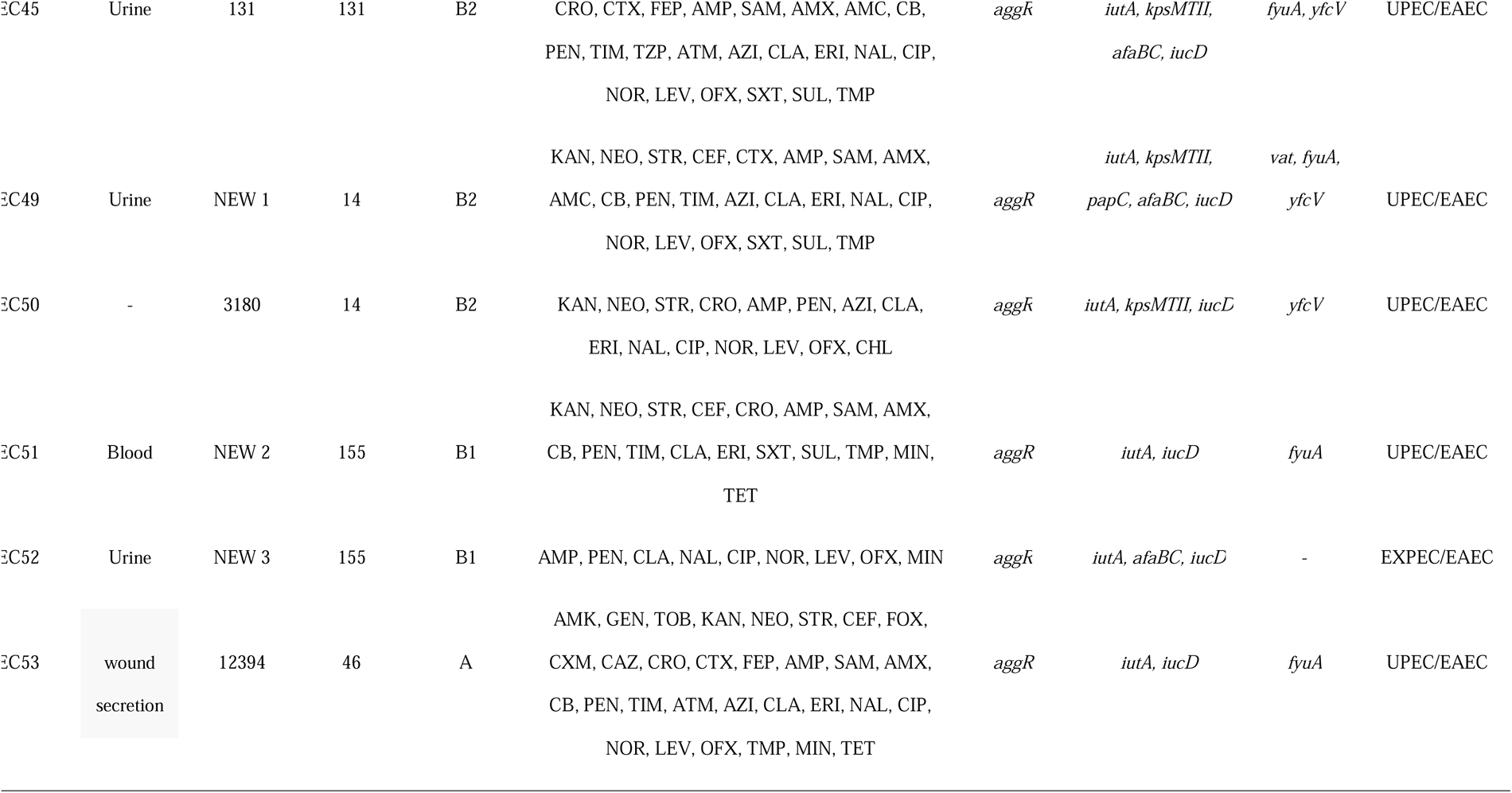
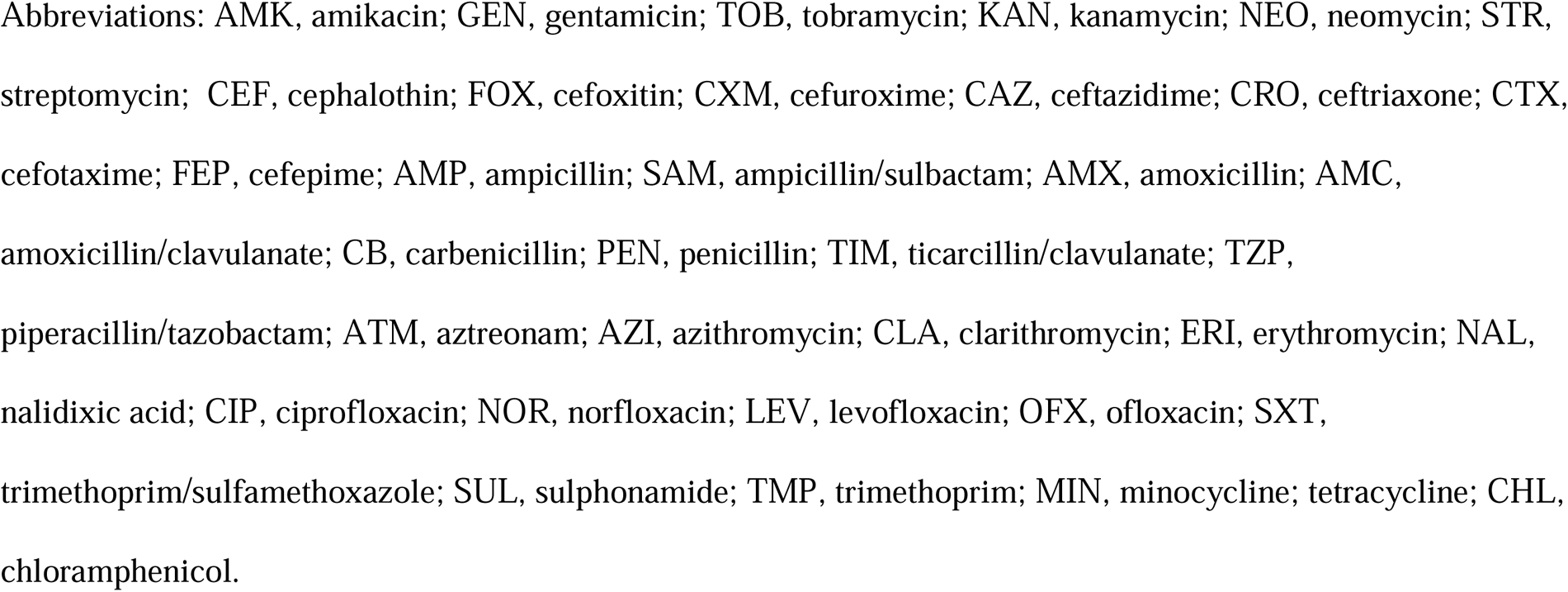
Clinical and genetic information of the hybrid *E. coli* strains in this study.

Considering the *E. coli* virulome, 9/19 virulence factors characterizing DEC and ExPEC pathotypes were found in the studied samples in different frequencies and combinations (Table 2). The presence of at least one extraintestinal virulence marker was observed in all isolates, corroborating the nature of the infection sites from which they were recovered. The *iucD* gene, which is part of the operon involved with aerobactin siderophore synthesis, was the most prevalent ExPEC marker among the isolates (12/15). This gene is associated with iron uptake systems, facilitating its survival and pathogenicity and playing a crucial role in the bacteria’s ability to cause infections and evade the host’s immune response. Among the 15 ExPEC isolates analysed, 12 (80%) had at least one marker used to define ExPEC and DEC pathotypes, being characterized as hybrid strains. The three strains classified as non-hybrids belonged to the pandemic lineages ST131/phylogroup B2 (EC44) and ST69/phylogroup D (EC39) and to the local ST8886 (EC40). Only 13 entries in Enterobase (https://enterobase.warwick.ac.uk/species/index/ecoli) have been found for ST8886, and in most cases, they corresponded to strains recovered from environmental sites in USA in 2015 and only one recovered from an animal source in Canada in 2022. Thus, this study reported the first clinical case belonging to this lineage.

Concerning the hybrid strains, it was verified the presence of UPEC/EAEC (n=10) and ExPEC/EAEC (n=2) (when UPEC markers were absent, or the strain had been recovered from a different body site other than urine). The *aggR* gene, considered a diarrheagenic factor that defines the EAEC pathotype, was found in all hybrid isolates, indicating that they would have the ability to cause intestinal disorders despite presenting in extraintestinal infections. EAEC is recognized as an important cause of diarrhoea worldwide and can be considered the most common DEC pathotype in *E. coli* in Brazil. However, *E. coli* strains with a hybrid enteroaggregative/uropathogenic (UPEC/EAEC) genotype have sporadically emerged as a cause of extraintestinal infections in Brazil [10–18]. Here we demonstrated the high prevalence of such hybrid pathotypes in distinct lineages causing infections in the Amazon region.

Most of UPEC/EAEC hybrid strains belonged to the pandemic and widespread ST131 (one of them was assigned to a new ST of the CC131) (Table 2). They were recovered from distinct extraintestinal sites and presented a heterogeneous profile of virulence markers. The unique non-hybrid ST131 strain was the EC44, which was a UPEC recovered from UTI that presented both UPEC and ExPEC virulence markers. On the other hand, local STs (ST9403, ST3180 and ST12394) belonging to different phylogroups (A, B1 and B2) and recovered from distinct body sites, including blood, were also classified as UPEC/EAEC hybrid strains.

Interestingly, in spite of the diversity of infection sites, most of the ExPEC strains were recovered from urine, and all but one were resistant to the tested fluoroquinolones (Table 2), which are the first choice antibiotics for treating UTIs. These findings probably accounted for treatment failure, compromising the success of clinical outcomes.

Interestingly, in spite of the UPEC/EAEC EC49 strain being a new local ST of CC14/phylogroup B2, this isolate exhibited the highest number of ExPEC and UPEC virulence markers, besides the EAEC marker *aggR* (Table 2). This demonstrates that not only pandemic high-risk clones display an enhanced pathogenicity and that emerging strains may possess a high level of virulence fitness.

The other class of hybrids (ExPEC/EAEC) corresponded to strains (n=2) that carried ExPEC virulence markers and the *aggR* EAEC marker. The strains belonged to a new ST from CC155/phylogroup B1 (EC52) and to the local ST1196 belonging to phylogroup AxB1 (EC47) (Table 2). A recent study in Mozambique demonstrated that a hybrid ExPEC/EAEC strain recovered from blood belonged to the pandemic ST131 lineage and presented a remarkable virulence profile [28]. These findings revealed that ExPEC/EAEC hybrids are associated with distinct lineages. Interestingly, both ExPEC/EAEC strains identified here were recovered from urine although no UPEC virulence marker had been identified. This was also the case of the non-hybrid EC39 strain, belonging to the pandemic ST69/phylogroup D, that had been characterized as a UPEC pathotype due to the site of infection (urine), although all UPEC virulence markers were absent (Table 2). These findings suggest that their ability to infect the urinary tract would be related to other mechanisms that were not investigated in this study. In fact, UPEC employs various virulence strategies, such as binding to urinary tract epithelial cells, iron acquisition, toxin production, and evasion of the host’s immune system. These virulence factors are commonly encoded by genes found in pathogenicity islands (PAIs), which are often mobilized through horizontal gene transfer and could have been lost from those UPEC strains [14]. Furthermore, the presence of UPEC factors in strains recovered from sites other than urine, such as the non-hybrid EC40 strain (Table 2), demonstrates their ability to colonize and survive in the urinary tract, but also that other virulence determinants were involved in the establishment of extraintestinal infections in those cases.

## CONCLUSION

This study contributes to a better understanding of the occurrence and pathogenic potential of hybrid strains of *E. coli*. Besides, the association of pathotype and lineage definition provides new information in the scenario of the global epidemiology of ExPEC hybrid strains.

## Data Availability

All data produced in the present work are contained in the manuscript

## Ethical statement

This study was approved by the Oswaldo Cruz Foundation Ethics Committee (CAAE: 39978114.5.0000.5248).

---

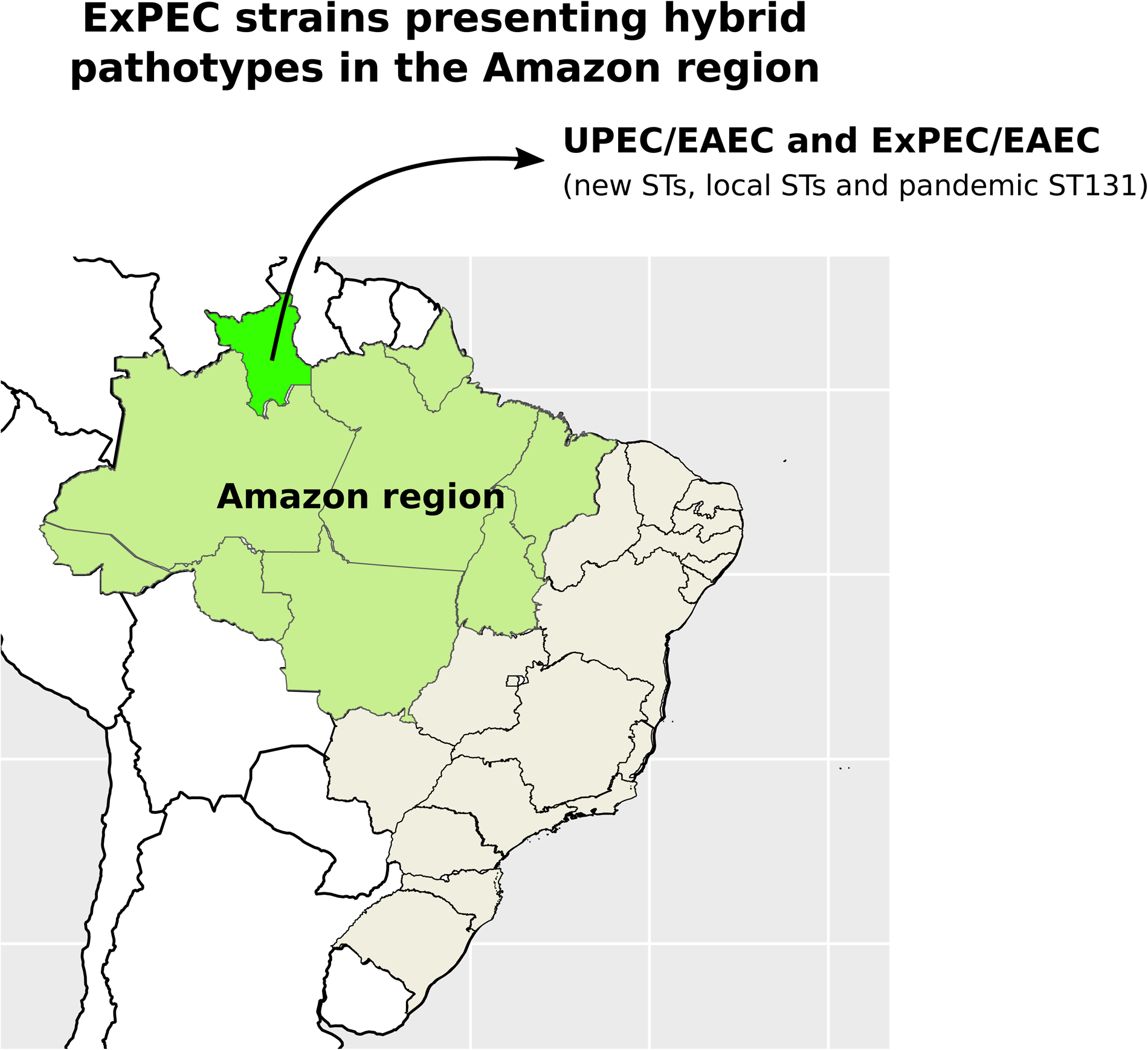

